# A rapid assessment of wastewater for genomic surveillance of SARS-CoV-2 variants at sewershed scale in Louisville, KY

**DOI:** 10.1101/2021.03.18.21253604

**Authors:** J. L. Fuqua, E.C. Rouchka, S. Waigel, K. Sokoloski, D. Chung, W. Zacharias, M. Zhang, J. Chariker, D. Talley, I. Santisteban, A. Varsani, S. Moyer, R. H. Holm, R. A. Yeager, T. Smith, A. Bhatnagar

**Affiliations:** Department of Pharmacology and Toxicology, University of Louisville, 505 S. Hancock St., Louisville, KY 40202, United States; Center for Predictive Medicine, University of Louisville, 505 S. Hancock St., Louisville, KY 40202, United States; Department of Computer Science and Engineering, University of Louisville, 522 East Gray St., Louisville, KY 40202, United States; KY-INBRE Bioinformatics Core, University of Louisville, 522 East Gray St., Louisville, KY 40202, United States; Department of Medicine, University of Louisville, 530 S. Hancock Jackson St., Louisville, KY 40402, United States; Department of Microbiology and Immunology, University of Louisville, 505 S. Hancock St., Louisville, KY 40202, United States; Department of Neuroscience Training, University of Louisville, 505 S. Hancock St, Louisville, KY 40202; Louisville/Jefferson County Metropolitan Sewer District, Morris Forman Water Quality Treatment Center, 4522 Algonquin Parkway, Louisville KY 40211, United States; The Biodesign Center of Fundamental and Applied Microbiomics, School of Life Sciences, Center for Evolution and Medicine, Arizona State University, Tempe, AZ 85287, United States; Department of Health Management and System Sciences, School of Public Health and Information Sciences, University of Louisville, 485 E. Gray St., Louisville, KY 40202, United States; Department of Public Health and Wellness, Louisville Metro Government, 400 E. Grays St., Louisville, KY 40202, United States; Christina Lee Brown Envirome Institute, University of Louisville, 302 E. Muhammad Ali Blvd., Louisville, KY 40202, United States; Department of Environmental and Occupational Health Sciences, School of Public Health and Information Sciences, University of Louisville, 485 E. Gray St., Louisville, KY 40202, United States

## Abstract

In this communication, we report on the genomic surveillance of SARS-CoV-2 using wastewater samples in Jefferson County, KY. In February 2021, we analyzed seven wastewater samples for SARS-CoV-2 genomic surveillance. Variants observed in smaller catchment areas, such as neighborhood manhole locations, were not necessarily consistent when compared to associated variant results in downstream treatment plants, suggesting catchment size or population could impact the ability to detect diversity.

---

The successful viral detection of severe acute respiratory syndrome coronavirus 2 (SARS-CoV-2) RNA in wastewater at various pooled scales (1-4) and discovery in the USA of B.1.1.7, B.1.351 and P.1 variants (5), has led to an interest in developing reliable population-level wastewater viral genomic surveillance.

The diversity of SARS-CoV-2 sequences reported to be circulating in the USA, have been determined by sequencing clinical samples; however, these variants can also be surveilled by sequencing wastewater samples (6-9). As of March 2021, the variants of concern -B.1.1.7, B.1.351, and P.1 have been widely detected in clinical samples from 47 states in the USA. In Kentucky, only five clinical cases have been linked to the presence of these variants (5),which could indicate incomplete surveillance. Broadening the application of genomic surveillance using wastewater in the community could enhance SARS-CoV-2 variant population monitoring.

In this communication, we report on the genomic surveillance of SARS-CoV-2 using wastewater samples in Jefferson County, KY. Samples were collected from manholes and treatment facilities, covering populations of 8,000 to 350,000 people (Table 1). RNA isolated from wastewater samples was used to quantify SARS-CoV-2 and analyze the genetic variation through high-throughput sequencing (See Supplementary Methods). Bioinformatics approaches were used to rapidly identify single nucleotide genetic alterations, which were compared with known variants of interest and concern.

**Table 1.** Summary of wastewater SARS-CoV-2 samples sequenced in this study, Louisville, KY.

| Sample ID | Sewershed population | Location | N1 (Ct) | Sequencing BWA Alignment Rate (%) |
| --- | --- | --- | --- | --- |
| **833** | 35,956 | Street line manhole leading to Treatment Plant #3<sup>a</sup> | 28 | 28.02 |
| **Treatment Plant #1** | 55,928 | Treatment Plant | 30 | 21.09 |
| **847** | 10,739 | Street line manhole leading to Treatment Plant #2 | 29 | 15.08 |
| **849** | 35,956 | Street line manhole leading to Treatment Plant #3<sup>a</sup> | 28 | 12.61 |
| **884** | 46,659 | Street line manhole leading to Treatment Plant #3<sup>a</sup> | 29 | 23.98 |
| **891** | 8,071 | Street line manhole leading to Treatment Plant #2 | 29 | 26.03 |
| **Treatment Plant #2** | 349,850 | Treatment Plant | 31 | 19.96 |

<sup>a</sup> Treatment Plant #3 samples had SARS-CoV-2 was detected but were below the threshold for individual mutations for review.

In February 2021, we analyzed seven wastewater samples for SARS-CoV-2 genomic surveillance (Figure 1). We did not detect genetic variations indicative of any current variant of concern, beyond the widespread D614G spike protein mutation (Supplementary Methods Tables 2-5). In all samples, we identified at least four of ten mutations consistent with the presence of the variant of interest B.1.429, and one sample contained seven of ten mutations (Table 2). The B.1.429 variant was confirmed in patient samples in Kentucky in January 2021 (10), and a single patient in the study area was reported to be positive for B.1.1.7 on February 9, 2021 (11). With our current metrics we flagged sites 833, 891, and Treatment plant #2 for potential presence of variant B.1.429 (3/7 sites). Differences in the scale of sample pooling in the community revealed unanticipated inconsistencies in variant representation. Specifically, variants observed in smaller catchment areas, such as neighborhood manhole locations, were not observed in downstream treatment plants, suggesting catchment size or population could impact the ability to detect diversity.

**Table 2.**
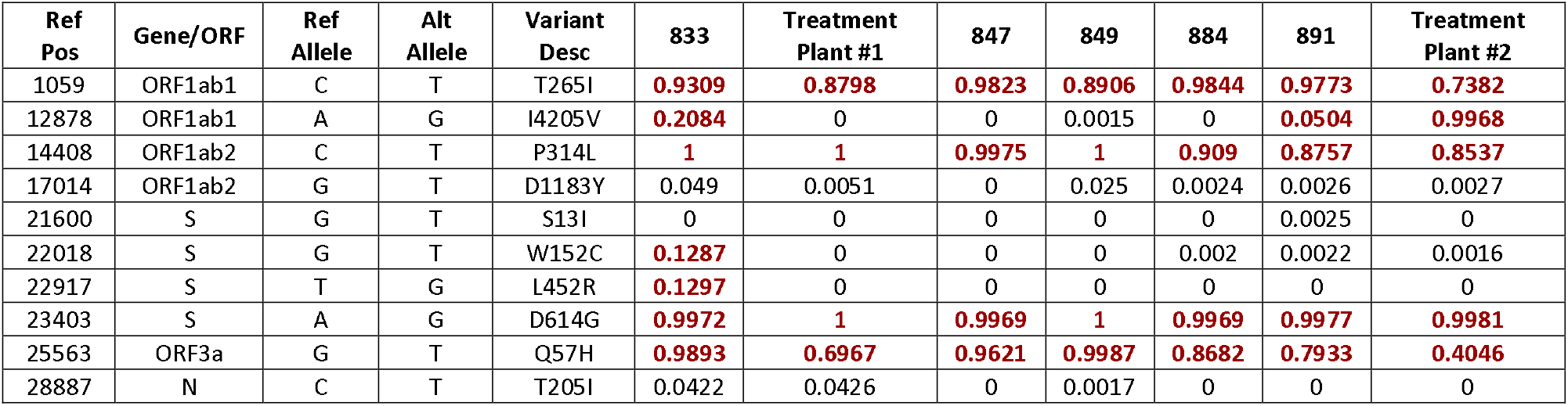
Summary of B.1.429 specific mutation prevalence by sample.

**Figure 1.**
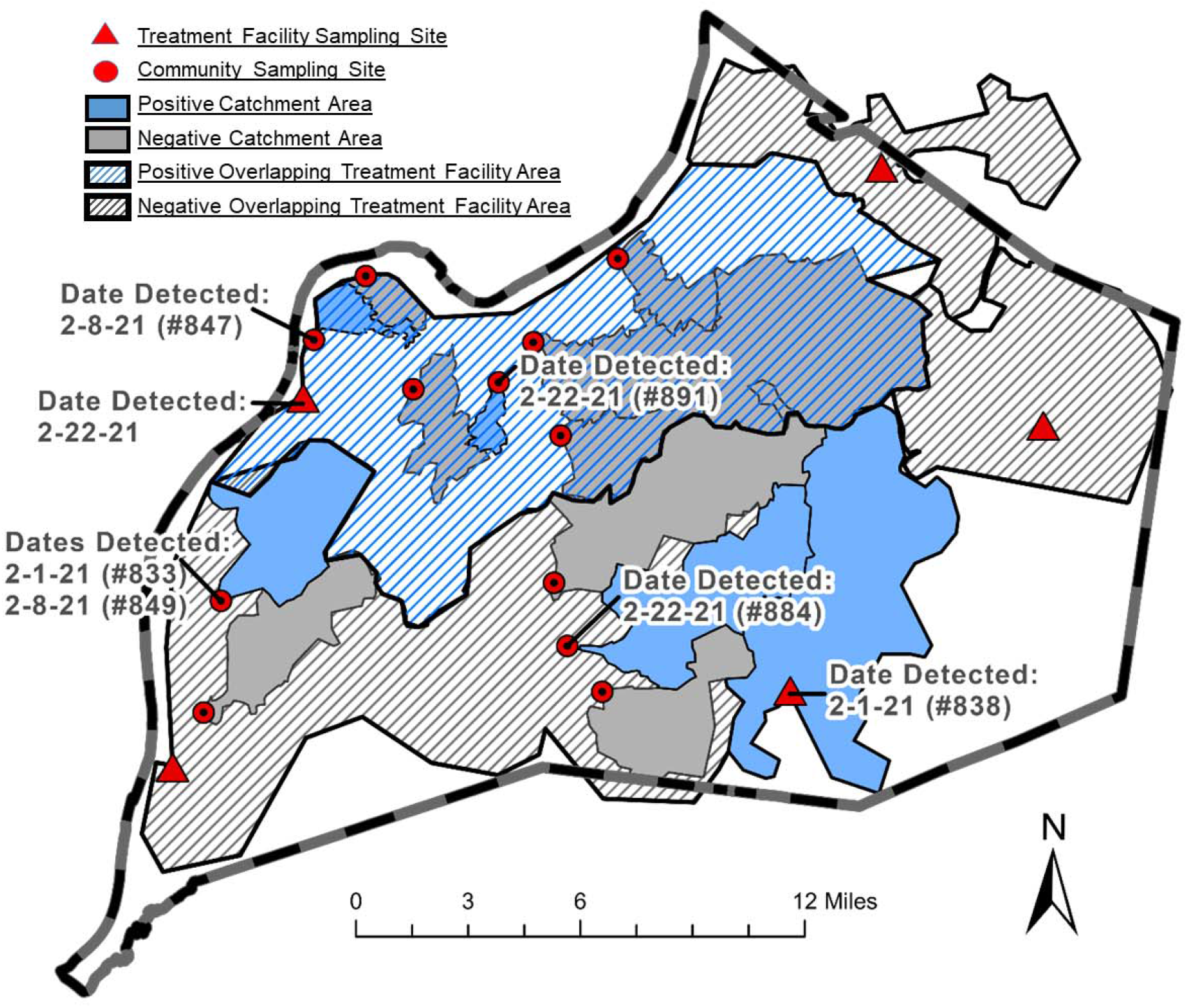
Study sites within Louisville, KY. Distribution of the sewershed area, treatment plants and community locations, in Jefferson County with corresponding dates, sampled. SARS-CoV-2 was detected at all sites. Samples that contained at least 50% of the single amino acid mutations for a variant with a nucleotide frequency above a 5 % threshold for individual mutations are flagged for review. This relatively low threshold serves the purpose of identifying geographic (sewershed) areas for heightened public health surveillance. With our current metrics we flagged sites 833, 891, and Treatment plant #2 for potential presence of variant B.1.429.

Given the highly variable viral genome sequence coverage recovered from wastewater samples, there is an urgent need to develop a set of consistent thresholds constituting positive/negative presence of a variant. Monitoring SARS-CoV-2 variants in wastewater may warn of an emerging variant of concern and identify variant dominance occurring when a new variant is introduced in a community. Wastewater genetic monitoring may be particularly useful in the context of limited clinical sample sequencing capacity because a broad perspective on the genetic diversity can be obtained from a few samples. To develop comprehensive epidemiological frameworks required to guide policy, population-level wastewater surveillance of viral genetic diversity should be complemented by clinical sample testing.

## Data Availability

The data is available from the Corresponding Author.

## Acknowledgments

This project was supported by funding from Louisville Metro Public Health and Wellness. Part of this work was performed with assistance of the UofL Genomics Facility, which was supported by NIH/NIGMS KY-INBRE P20GM103436, the J.G. Brown Cancer Center, University of Louisville, and user fees. The following reagent was deposited by the Centers for Disease Control and Prevention and obtained through BEI Resources, NIAID, NIH: Quantitative PCR (qPCR) Control RNA from Heat-Inactivated SARS-Related Coronavirus 2, Isolate USA-WA1/2020, NR 52347.

## Ethics

The University of Louisville Institutional Review Board classified this project as Non-Human Subjects Research (NHSR) (reference #: 717950).

## Supplementary Methods

### Wastewater sample Prep

Wastewater samples were collected on February 1, 8 and 22, 2021. In brief, a 24-hour composite raw wastewater sample was collected into a sterile 125ml polyethylene terephthalate bottle. Viral particles where concentrated using PEG precipitation methods. For each sample, 40ml of chilled wastewater was passed through a 70 µm cell strainer and PEG 8000 and (0.5g) NaCl were added to a final concentration of 12.5 mM and 210 mM, respectively. Samples were refrigerated overnight at 4°C and then centrifuged at 16,000 x g for 30mins at 4°C. The pellet was resuspended with 1.1ml TRIzol (Thermo Scientific # 15596018) and transferred to a sterile microfuge tube. The TRIzol sample was then incubated for 5 mins at room temperature and then centrifuged at 12,000 x g for 5 min at 4°C. The sample was then divided into two 500µl samples, one for isolation and one for archiving at -80°C. The sample for isolation had an additional 500µl of TRIzol added and 900µl of 100% Ethanol. Samples were vortexed and the RNA was isolated using a Direct-zol™ 96 MagBead RNA kit (Zymo Research, R2102) with RNA eluted in 100µl of DNAse/RNAse Free Water. RNA cleanup was done using the RNeasy® PowerClean® Pro Cleanup Kit (Qiagen #13995-50) according to the manufacturer’s instructions with RNA eluted in 60µl of DNAse/RNAse Free Water. Purified RNA was inspected for yield and quality using a NanoDrop 1000. Number of viral copies in each sample was determined using a probe-based RT-qPCR on a QuantStudio 3 (Applied Biosystems) real-time PCR system using Taq 1-Step Multiplex Master Mix (Thermo Fisher #A28527). The primer and probe sequences are shown in Table 1 with 5 primer/probe sets used for each sample and all samples ran in triplicate. 4µl of sample was used for each 20µl reaction. PCR cycling conditions were 25°C for 2 min, 50°C for 10mins, 95°C for 2 min and 45 cycles of 95°C for 2 sec and 60°C for 30 sec. We generated a standard curve for each primer-probe set used and fit the Ct values to extrapolate copies per mL of wastewater. For this publication we are only reporting on the N1 Ct values generated from this methodology.

### cDNA Synthesis

The Superscript® IV First-Strand Synthesis System (Thermo Fisher #18091050) was used to generate cDNA with random hexamer primers. The RT reaction was mixed according to manufacturer’s instructions with a final reaction volume of 20 µl and 5 µl of our template RNA added to the mixture. The reverse transcriptase incubation step was performed with sequential incubation at 23°C for 10 min, 50°C for 30 min, and 80°C for 10 min, according to the manufacturer’s protocol with adjustment of the incubation times recommended by Swift Biosciences SNAP low input protocol.

### Library Prep

Libraries were prepared using the Swift Biosciences SNAP low input protocol for SARS-CoV-2 (Swift Bioscience, Ann Arbor, MI, Cat # COSG1V2-96, SN-5×296). 10 µl of cDNA was combined with 20µl of reaction mix and proceeded with multiplex PCR according to protocol. The PCR product was cleaned up using SPRIselect beads (Beckman Coulter, Brea, CA, Cat. No. B23318) at a 1.0X ratio. The purified sample/beads mix was resuspended in 17.4 µl of TE buffer provided in the post-PCR kit. Samples were indexed through PCR with the SNAP Unique Dual Indexing Primers (Swift Bioscience, Ann Arbor, Cat. # SN91096-1-PLATE). The indexing PCR product was further cleaned up and eluted from the beads using a 0.65X PEG NaCl clean-up. The purified libraries were then eluted in 22µL of TE buffer and transferred to fresh tubes and stored at -20°C. For some of the samples (884, 891, and Treatment Plant #2), 1 additional cycle was added to the multiplex PCR and 2 additional cycles were added to the indexing PCR to obtain higher library yields. The library concentration was measured using the Qubit dsDNA HS Assay Kit (Thermo Fisher, Waltham, MA, Q32851). The libraries’ size distribution was checked on the Agilent Bioanalyzer using the DNA High Sensitivity Kit (Agilent Technologies, Cat# 5067-4626). Library normalization was performed according to SwiftBio’s Normalase 2nM final pool protocol. 5 µl of Normalase I Master Mix were added to each 20µl library eluate for a final pool of 2nM and thoroughly mixed. Samples were placed in the thermocycler to incubate at 30°C for 15 min. 5 µl of each library were pooled, and 1µl of Normalase II Master Mix per library was added and thoroughly mixed. The library pool was placed in the thermocycler to incubate at 37°C for 15 min. 0.2 μl of Reagent X1 per library was added to the pool to inactivate Normalase II at 95°C for 2 min and held at 4°C.

### Sequencing

Library pool and PhiX were denatured and diluted following Illumina’s directions. Libraries with 1% PhiX spike-in were sequenced at read length 2 x 150 bp using the MiSeq Reagent Kit v2 300 cycle (Illumina, San Diego, CA, Cat# MS-102-2002), or the NextSeq 500/550 Mid Output Kit v2.5 300 Cycles (Illumina, San Diego, CA, Cat# 20024905), targeting 1-5 M reads per library.

### Data analysis

Sequencing reads were analyzed using a custom bioinformatics pipeline. Low quality bases were trimmed using Trimmomatic v0.38 (1), and were then aligned to the NC_045512.2 reference genome using bwa mem v 0.7.17-r1188 (2). Single nucleotide variants (SNVs) relative to the reference were detected using bcftools mpileup (3). SNVs occurring in at least 5% of the reads with at least five separate supporting instances were marked for further interrogation. SNVs occurring at locations of interest as they relate to specific SARS-CoV-2 variants (B.1.1.7, B.1.351, B.1.526, P.1, and B.1.429) were reported for all of the samples (Supplementary Methods Tables 2-5).

**Table 1.**
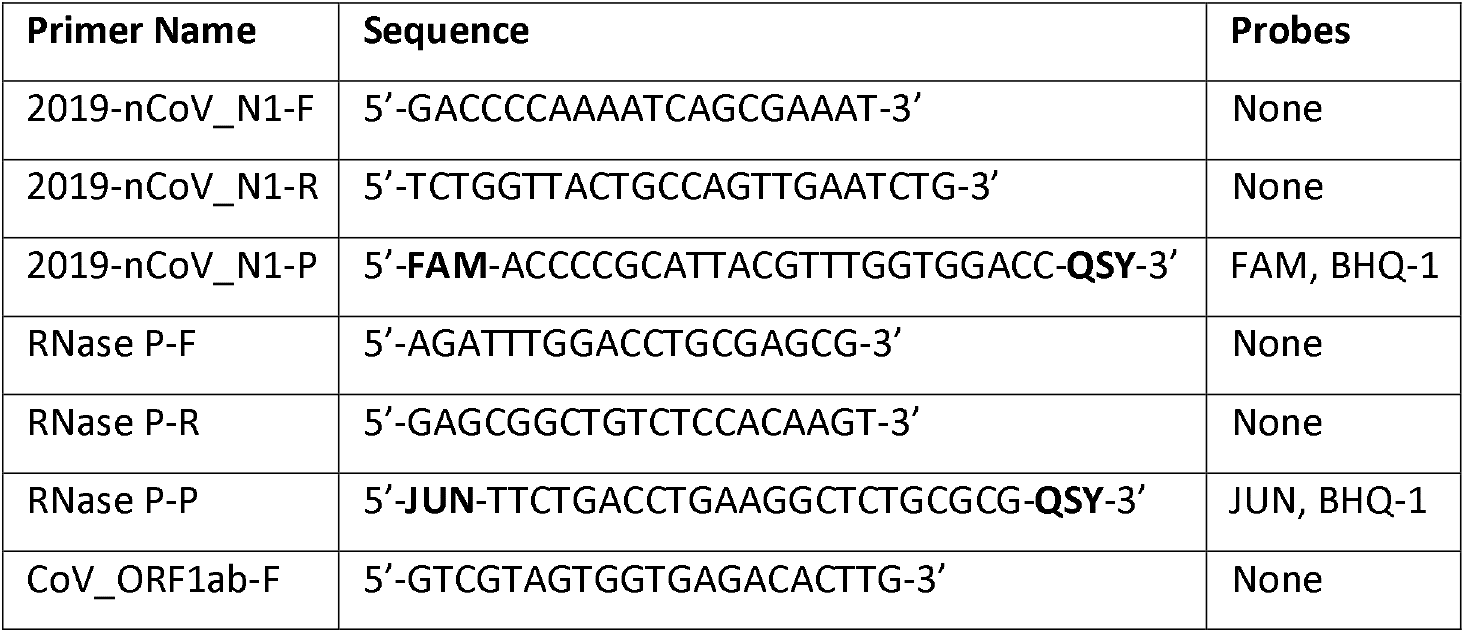
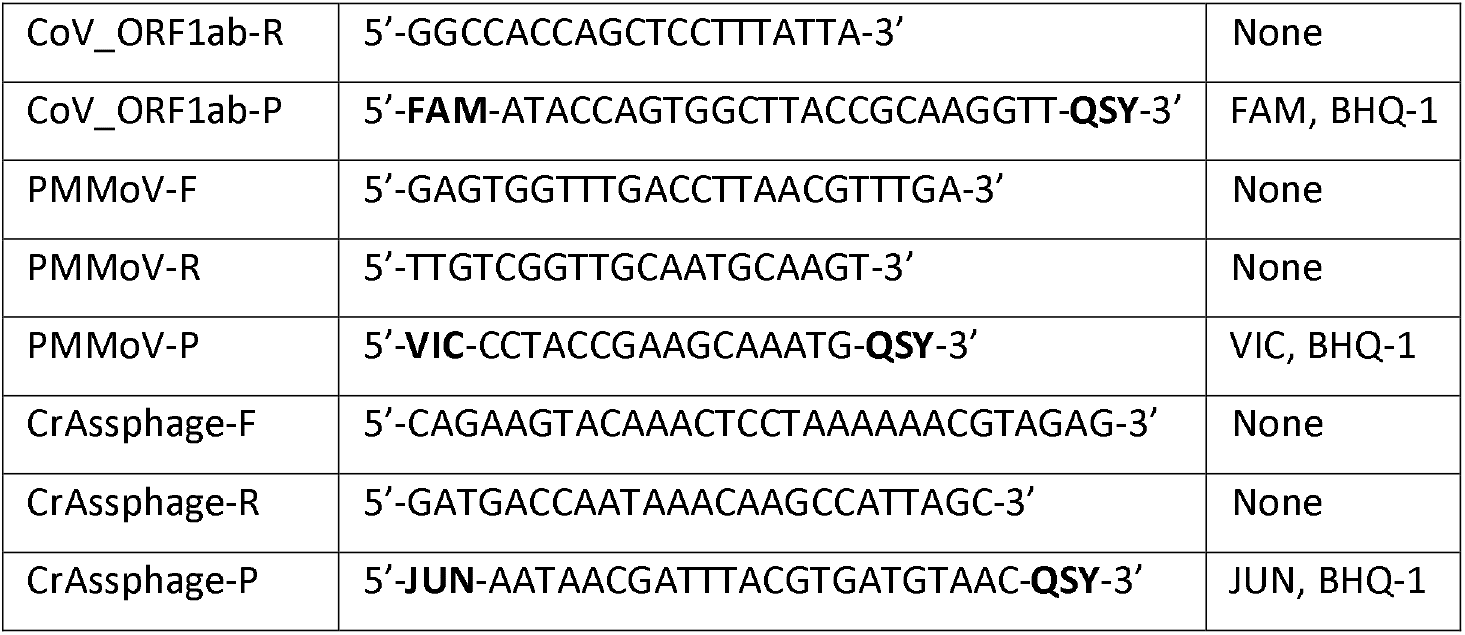
Primer and probe sequences used for RT-qPCR

**Table 2.**
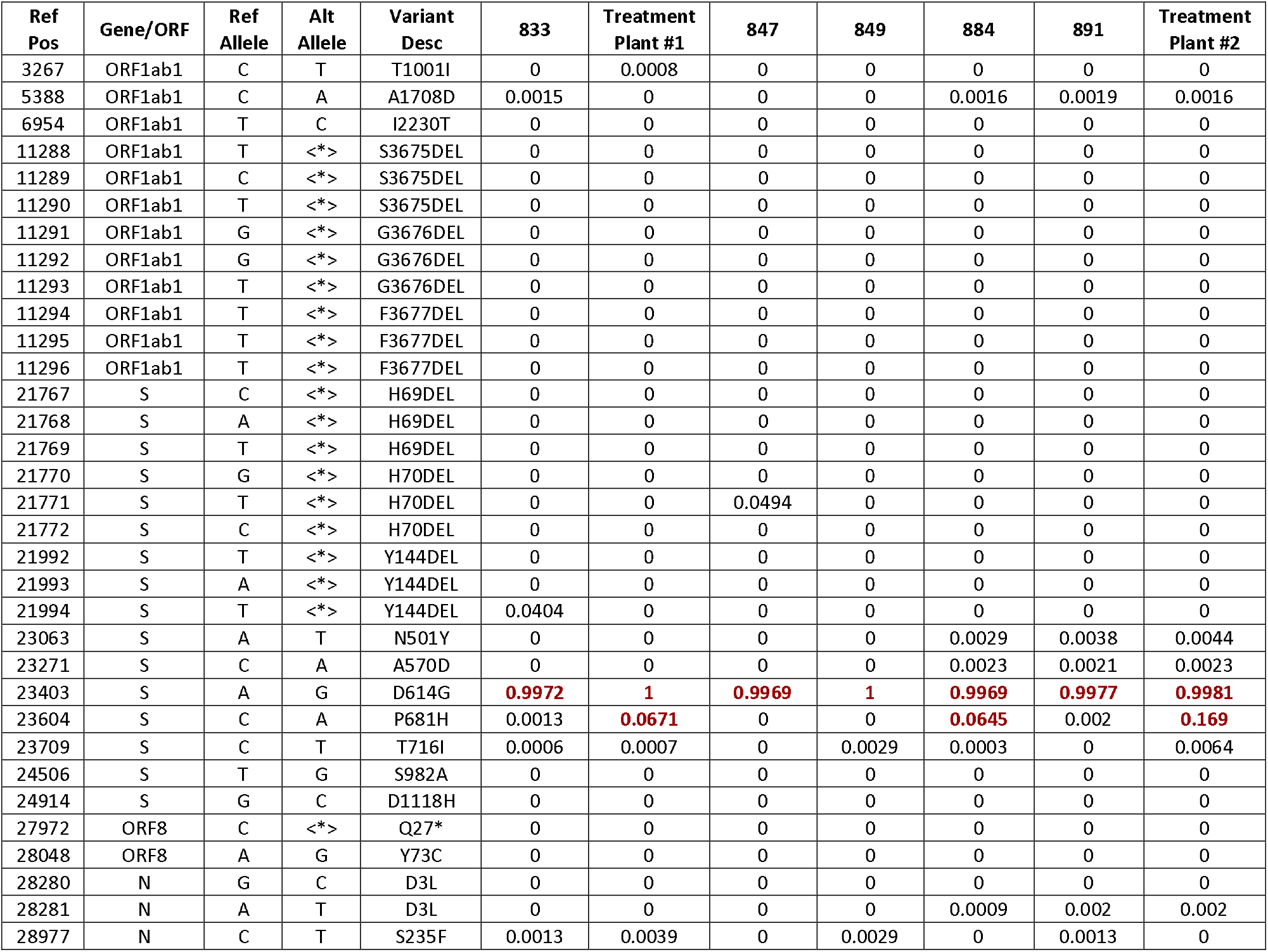
Summary of B.1.1.7 specific mutation prevalence by sample

**Table 3.** Summary of B.1.351 specific mutation prevalence by sample

| Ref Pos | Gene/ORF | Ref Allele | Alt Allele | Variant Desc | 833 | Treatment Plant #1 | 847 | 849 | 884 | 891 | Treatment Plant #2 |
| --- | --- | --- | --- | --- | --- | --- | --- | --- | --- | --- | --- |
| 5230 | ORF1ab1 | G | T | K1655N | 0.0006 | 0 | 0 | 0.0014 | 0.0023 | 0.0022 | 0.0021 |
| 22813 | S | G | C | K417N | 0 | 0 | 0 | 0 | 0 | 0 | 0 |
| 23012 | S | G | A | E484K | 0 | 0 | 0 | 0 | 0 | 0 | 0 |
| 23063 | S | A | T | N501Y | 0 | 0 | 0 | 0 | 0.0029 | 0.0038 | 0.0044 |
| 23403 | S | A | G | D614G | **0.9972** | **1** | **0.9969** | **1** | **0.9969** | **0.9977** | **0.9981** |
| 23664 | S | C | T | A701V | 0 | 0.0018 | 0.0051 | 0.0035 | 0.0009 | 0 | 0 |
| 26456 | E | C | T | P71L | 0 | 0.0069 | 0 | 0 | 0 | 0 | 0 |
| 28887 | N | C | T | T205I | 0.0422 | 0.0426 | 0 | 0.0017 | 0 | 0 | 0 |

**Table 4.** Summary of B.1.526 specific mutation prevalence by sample

| Ref Pos | Gene/ORF | Ref Allele | Alt Allele | Variant Desc | 833 | Treatment Plant #1 | 847 | 849 | 884 | 891 | Treatment Plant #2 |
| --- | --- | --- | --- | --- | --- | --- | --- | --- | --- | --- | --- |
| 21575 | S | C | T | L5F | 0 | 0 | 0.0051 | 0 | 0 | 0.011 | 0 |
| 21846 | S | C | T | T95I | 0 | 0 | 0 | 0 | 0 | 0 | 0 |
| 22320 | S | A | G | D253G | 0 | 0 | 0 | 0 | 0 | 0 | 0 |
| 23012 | S | G | A | E484K | 0 | 0 | 0 | 0 | 0 | 0 | 0 |
| 23403 | S | A | G | D614G | **0.9972** | **1** | **0.9969** | **1** | **0.9969** | **0.9977** | **0.9981** |
| 23664 | S | C | T | A701V | 0 | 0.0018 | 0.0051 | 0.0035 | 0.0009 | 0 | 0 |

**Table 5.**
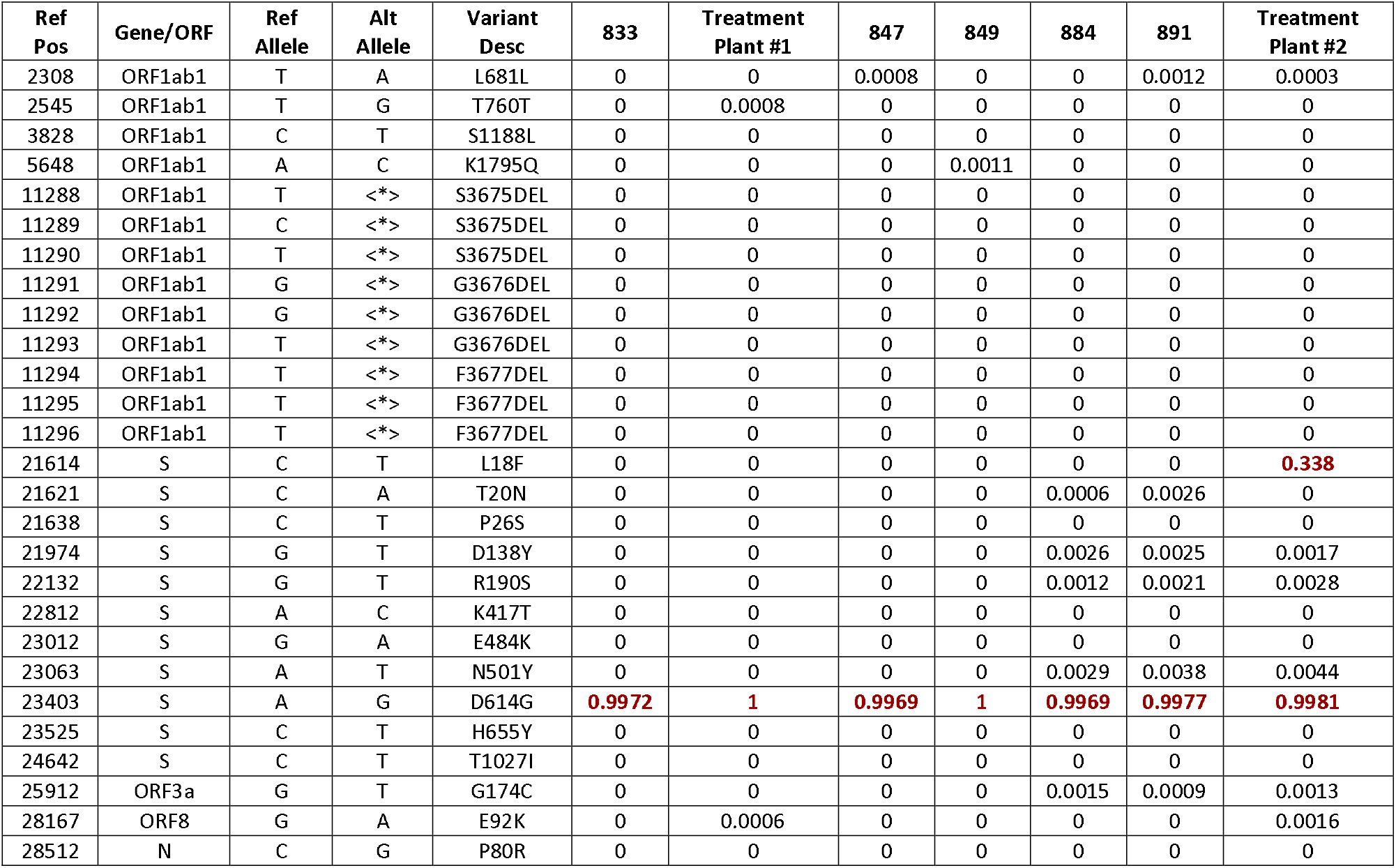
Summary of P.1 specific mutation prevalence by sample

